# Are We Falling Short? Evaluating the Accuracy of Common Clinical Fall Risk Assessments in Stroke Survivors: A Systematic Review and Meta-Analysis

**DOI:** 10.1101/2025.06.15.25329158

**Authors:** Marina Meyer-Vega, Nojan Valadi, Daniel J. Goble, Niyati Baweja, Harsimran S. Baweja

## Abstract

Stroke survivors experience a significant risk of falls, with 73% falling within the first year post stroke. Clinical practice guidelines currently use more than 27 assessment tools to evaluate fall risk in this population. However, there are conflicting findings regarding their diagnostic accuracy in correctly identifying those at risk of falling. This systematic review and meta-analysis evaluates the sensitivity and specificity of the Timed Up and Go and the Berg Balance Scale, which are the most commonly used clinical assessments for identifying fall risk among stroke survivors. Our data search included Web of Science, PubMed, and Ovid Medline. This protocol was registered in PROSPERO (CRD420251004460) prior to data extraction. Fifteen studies, comprising 1,492 stroke survivors, were included in the meta-analysis. We found a negative association between these tests and their ability to accurately identify fall risk in this population (OR = 0.469, 95% CI [-0.230, 1.169], however, this was not statistically significant (*p* = 0.188). No heterogeneity was observed across studies (τ^2^ = 0.000; I² = 0%). We found considerable variability in cut-off values across protocols, without significant moderating effects of these thresholds on their diagnostic accuracy. No publication bias was detected according to the Egger’s weighted test (t (20) = 0.024, *p* = 0.981) and the rank correlation test (τ = - 0.030, *p* = 0.867). Future research should focus on developing or implementing an objective stroke-specific fall risk assessment with appropriate cut-off values that better capture the underlying mechanisms of fall risk in stroke survivors to improve fall prevention strategies and rehabilitation care.

## INTRODUCTION

Stroke is a major global health concern, affecting approximately 15 million people worldwide each year [1]. In the United States alone, every 40 seconds, someone suffers from a stroke, making it one of the leading causes of death and long-term disability in the country [2]. The neurological damage from stroke often results in multiple stroke-related impairments that significantly contribute to reduced balance and fall risk, including neuromuscular weakness, lack of multisensory integration, reduced attention, and deficits in vision and spatial awareness [3–7]. Consequently, 7% of stroke survivors experience a fall in the first week and 73% in the first year post-stroke [3, 8].

In light of the above, stroke survivors are six times more likely to experience a fall when compared with healthy individuals [9, 10]. Falls can lead to serious consequences, including fractures, reduced quality of life, prolonged length of hospital stays, and a heavy financial burden [11, 12]. Moreover, individuals with a history of falls are more susceptible to future falls and increased mortality [13, 14]. This makes fall prevention a significant economic and healthcare priority in this population, highlighting the importance of clinical assessment tools and tests to specifically evaluate fall risk in stroke survivors, which are critical for high-quality, effective rehabilitation and preventing future falls [15, 16].

Clinical assessment tools have the purpose of providing objective and reproducible data that can inform diagnosis and treatment planning, predict outcomes and/or prognosis, and monitor individual progress over time [16–19]. Despite these clear objectives, clinicians currently use more than 27 different tests, tools, and instruments to assess fall risk in stroke survivors [16]. This overabundance and inconsistency present a challenge to correctly inform clinical practice guidelines (CPGs). It complicates decision-making when selecting appropriate assessment tools. Moreover, standardized assessments are an essential element of evidence-based rehabilitation [20]. However, in this case, the lack of standardization across tests complicates the comparison of outcomes and identifying the most effective intervention [21].

In 2019, Dos Santos et al. reviewed the most common tools used in current CPGs for stroke survivors worldwide [16]. Among the 19 CPGs included in the review, the most frequently used were the Timed Up and Go (80%), the Berg Balance Scale (90%), the 6-Minute Walk Test (80%), and the 10-Meter Walk Test (70%) [16]. However, some groups have raised concerns regarding the diagnostic accuracy of these tests, specifically their sensitivity and specificity in differentiating between fallers and non-fallers, suggesting that clinicians should exercise caution in their application [22, 23].

Sensitivity refers to a test’s ability to correctly identify those at risk of falling (true positives), while specificity refers to a test’s ability to correctly identify those not at risk of falling (true negatives) [24, 25]. Findings suggest that common stroke assessment tools may overestimate or miscategorize fall risk [23, 26]. Categorizing patients as high fall risk when they are not leads to unwarranted additional assessments and interventions [27]. Perhaps more concerning, misclassification can create an elevated fear of falling, potentially decreasing their independence and discouraging them from engaging in activities they are fully capable of performing [28].

In contrast, other groups have shown that common stroke assessment tools have excellent interrater reliability [29–31]. They are easy to administer and serve as an effective clinical tool for assessing functional mobility and balance [32, 33]. Given the conflicting findings on the sensitivity and specificity of the most commonly used fall risk assessment tools and their critical role in fall prevention strategies for stroke survivors, a more comprehensive evaluation of their diagnostic efficacy is necessary.

This systematic review and meta-analysis aim to evaluate the sensitivity and specificity of the most commonly used clinical assessments for identifying fall risk among stroke survivors. The findings of this study will: 1) inform current CPGs whether existing tests demonstrate sufficient diagnostic accuracy to predict fall risk and guide rehabilitation interventions and outcomes accurately; 2) determine which of these assessment tools demonstrate the highest sensitivity and specificity, potentially establishing it as a standard across CPGs; and 3) if no current assessment tools demonstrate adequate diagnostic accuracy for predicting fall risk in this population, highlight the need to develop and implement an objective, sensitive, stroke-specific fall risk assessment tool to improve diagnostic accuracy and its impact on the rehabilitation outcomes in stroke survivors.

## METHODS

A systematic search was conducted according to the updated Preferred Reporting Items for Systematic Reviews and Meta-Analysis (PRISMA) guidelines [34]. The protocol was registered in the International Prospective Register of Systematic Reviews (https://www.crd.york.ac.uk/PROSPERO/; Unique identifier: CRD420251004460).

### Search Strategy and Data Extraction

An electronic search was performed by the lead author (MMV) in several electronic databases: Web of Science, PubMed, SportDiscuss, and Ovid MEDLINE from inception up to 03 March 2025. The search terms were prepared by the lead author (MMV) using Boolean operators in the following format: ((Stroke OR Cerebrovascular OR CVA) AND (Accidental fall* OR fall* OR fall risk OR fall prevention OR fall assessment*) AND (postural sway OR postural control OR postural balance OR dynamic balance OR static balance) AND (risk assessment OR balance test OR balance assessment OR balance measure OR clinical measure* OR Timed Up and Go OR TUG OR Berg Balance OR BBS OR 6 Minute Walk Test OR 6MWT OR 10 Meter Walk Test or 10MWT) AND (sensitivity OR specificity OR measurement properties OR psychometric OR predictive value OR multivariate analysis)).

Two reviewers independently screened the titles and abstracts of all remaining studies for eligibility. After removing studies irrelevant to the research question, full-text articles of potentially eligible studies underwent a second independent screening. Any disagreements between reviewers were resolved through discussion and consensus. This systematic review and meta-analysis included studies that met the following criteria. *Inclusion and Exclusion Criteria*

Studies were included if they (a) were articles published in English or Spanish; (b) Participants with a medical diagnosis of stroke without any other neurological or musculoskeletal condition; (c) included the outcome of at least one of the following tests: (1) Timed Up and Go; (2) Berg Balance Scale; (3) 10-Meter Walk Test; (4) 6-Minute Walk Test; and (d) provided sufficient data to calculate effect size (e.g., number of true fallers, number of true non-fallers, sensitivity and specificity of the test). Studies classified as case reports, articles presented at conferences, systematic reviews, or meta-analysis studies were excluded from the analysis.

### Study Selection

The lead author (MMV) systematically extracted relevant information from all included studies using Microsoft Excel (Microsoft Corporation, Redmond, WA, USA, version 16.88). The data extracted included authors, year of publication, study design, sample size (total fallers and non-fallers), diagnostic performance metrics (sensitivity and specificity, true positives (TP), true negatives (TN), false positives (FP), and false negatives (FN)), and participants’ demographics (e.g., affected side, sex, and age).

For studies that did not directly report the contingency table values (TP, TN, FP, FN), we derived these values from the reported sensitivity, specificity, and the known number of true fallers and non-fallers. TP were calculated as the product of sensitivity and the total number of fallers. TN were calculated as the product of specificity and the total number of non-fallers. FP were derived by multiplying the complement of specificity (1-specificity) by the total number of non-fallers. Finally, FN were calculated by subtracting the TP from the total number of fallers [35].

### Statistical Analysis

The effect size for each study was calculated using the natural logarithm of odds ratio (logOR), along with its variance using the following formulas [36, 37]:

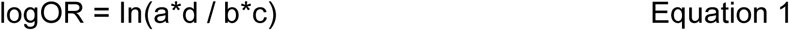

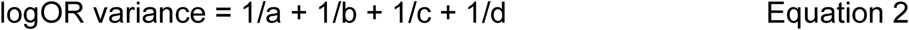

Where a represents the number of true positives, d represents the number of true negatives, b represents the number of false positives, and c represents the number of false negatives. To facilitate the interpretation of the results, logOR and its confidence intervals were back transformed into OR for all interpretations using the following formulas [38]:

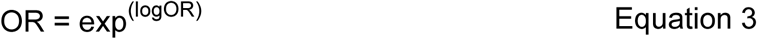

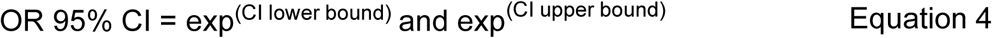

An OR of more than 1 indicates that the clinical tests effectively discriminate between fallers and non-fallers. The higher the OR the stronger the clinical test’s ability to correctly classify stroke survivors.

This meta-analysis used a weighted random-effects model with the restricted maximum likelihood estimator (REML) [39]. The heterogeneity across included studies was quantified using the estimated tau-squared (τ^2^) value, representing the variance of the true effect sizes [40]. The I^2^ statistic was also calculated to determine the percentage of total variability attributable to heterogeneity [40, 41]. A Q-test was conducted to assess the presence of heterogeneity, with its associated p-value indicating whether the observed heterogeneity was statistically significant [40, 41]. Additionally, we performed a subgroup analysis to compare the diagnostic accuracy between the tests and a meta-regression analysis to examine whether the cut-off values used for each test significantly moderate their diagnostic accuracy. The model results were reported as the estimated effect size (OR), standard error (SE), z-value, and p-value. The 95% confidence interval (CI) for the effect size was calculated to provide a range of plausible values. All statistics were executed using the statistical software package JASP (JASP Team, University of Amsterdam, Netherlands, version 0.19.2).

### Sensitivity Analysis and Risk of Bias

We evaluated publication bias using Egger’s weighted regression test. Moreover, funnel plot asymmetry was assessed through a rank correlation test. To determine the robustness of the results, we conducted a leave-one-out sensitivity analysis by systematically removing each study individually from repeated meta-analyses to evaluate its impact on the overall findings.

## RESULTS

### Search Results

Our search strategy identified 899 studies, with 674 remaining after removing 225 duplicates. Following title and abstract screening, 138 studies were selected for full-text review. Of the remaining studies, only 15 were eligible for data extraction, as detailed in Fig. 1. Of these 15 studies, six exclusively evaluated the Berg Balance Scale [42–47], three focused on the Timed Up and Go [48–50], and six examined both tests (Table 1) [51–56]. Furthermore, our search identified only three eligible studies for the 6-Minute and 10-Meter Walk Test. This small sample size was insufficient to calculate their effect size. Consequently, we excluded all studies evaluating only these tests from our final analysis.

**Figure 1.**
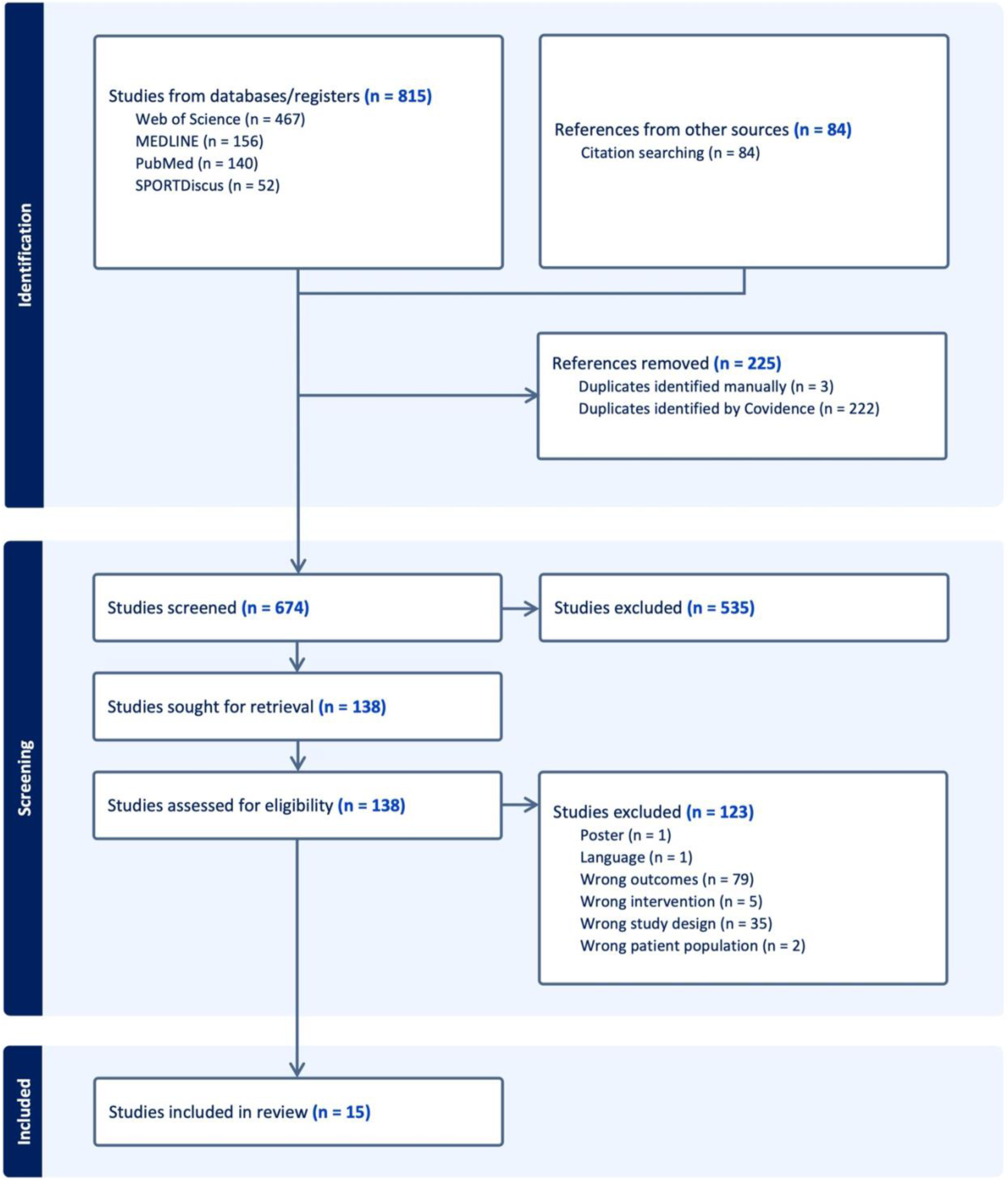
PRISMA 2020 flowchart of study selection.

**Table 1.**
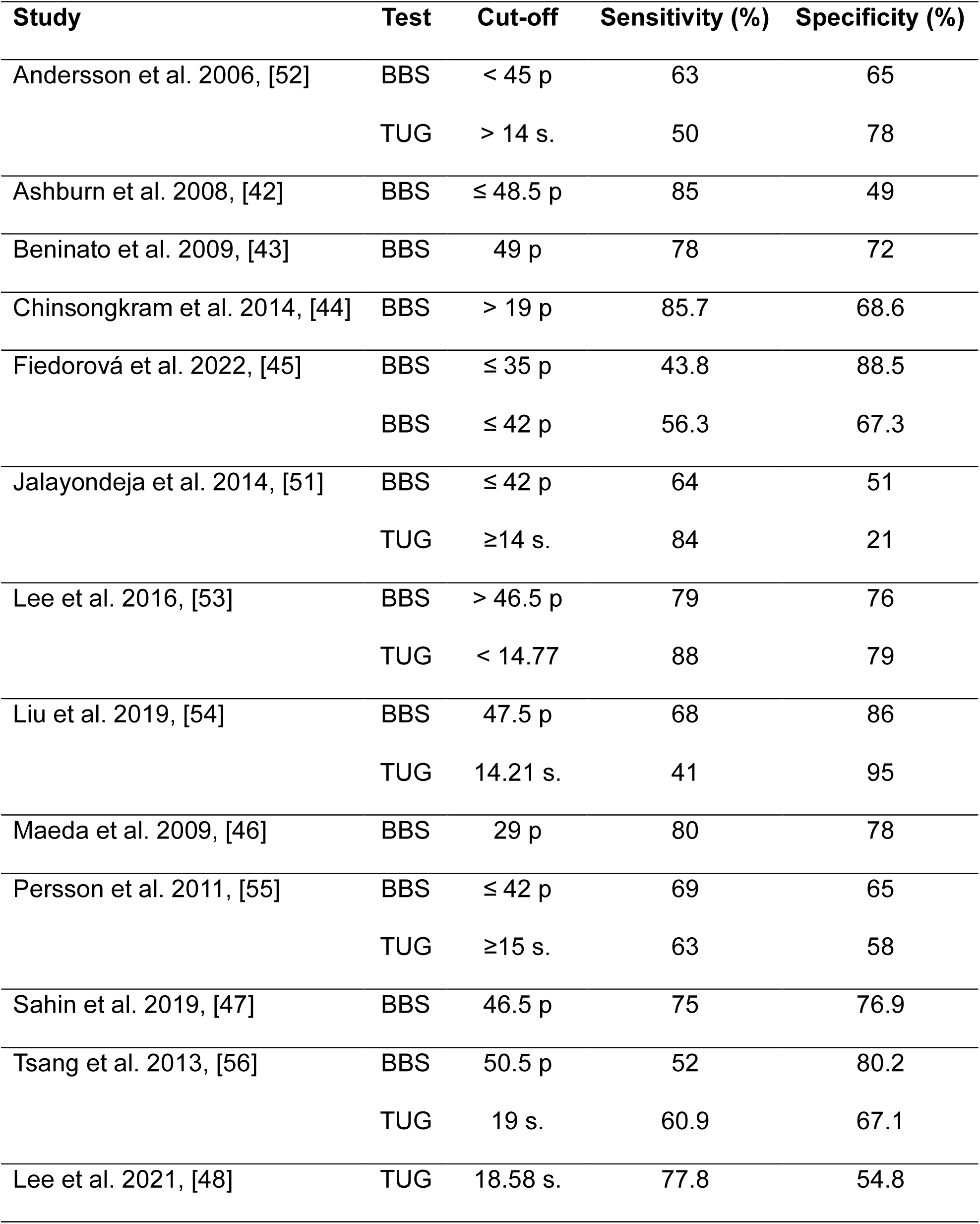
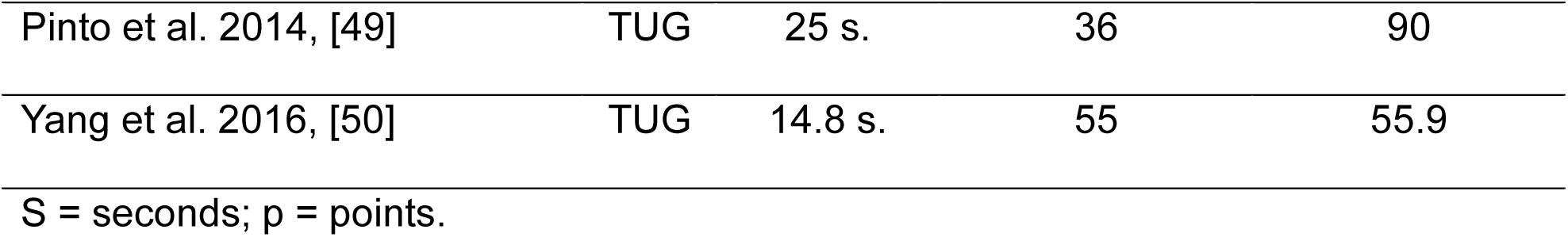
Characteristics of the Berg Balance Scale (BBS) and the Timed Up and Go (TUG) test from the 15 included studies.

| Study | Test | Cut-off | Sensitivity (%) | Specificity (%) |
| --- | --- | --- | --- | --- |
| Andersson et al. 2006, [52] | BBS | < 45 p | 63 | 65 |
|  | TUG | > 14 s. | 50 | 78 |
| Ashburn et al. 2008, [42] | BBS | ≤ 48.5 p | 85 | 49 |
| Beninato et al. 2009, [43] | BBS | 49 p | 78 | 72 |
| Chinsongkram et al. 2014, [44] | BBS | > 19 p | 85.7 | 68.6 |
| Fiedorová et al. 2022, [45] | BBS | ≤ 35 p | 43.8 | 88.5 |
|  | BBS | ≤ 42 p | 56.3 | 67.3 |
| Jalayondeja et al. 2014, [51] | BBS | ≤ 42 p | 64 | 51 |
|  | TUG | ≥14 s. | 84 | 21 |
| Lee et al. 2016, [53] | BBS | > 46.5 p | 79 | 76 |
|  | TUG | < 14.77 | 88 | 79 |
| Liu et al. 2019, [54] | BBS | 47.5 p | 68 | 86 |
|  | TUG | 14.21 s. | 41 | 95 |
| Maeda et al. 2009, [46] | BBS | 29 p | 80 | 78 |
| Persson et al. 2011, [55] | BBS | ≤ 42 p | 69 | 65 |
|  | TUG | ≥15 s. | 63 | 58 |
| Sahin et al. 2019, [47] | BBS | 46.5 p | 75 | 76.9 |
| Tsang et al. 2013, [56] | BBS | 50.5 p | 52 | 80.2 |
|  | TUG | 19 s. | 60.9 | 67.1 |
| Lee et al. 2021, [48] | TUG | 18.58 s. | 77.8 | 54.8 |
| Pinto et al. 2014, [49] | TUG | 25 s. | 36 | 90 |
| Yang et al. 2016, [50] | TUG | 14.8 s. | 55 | 55.9 |

S = seconds; p = points.

### Study Characteristics

This systematic review analyzed data from 1,492 stroke survivors across the 15 included studies (Table 2). Among participants, 35.8% (534) experienced falls, and 64.2% (958) were non-fallers. The demographic and clinical characteristics presented are based solely on variables that were reported in the studies. Participants had a mean age of 58±6 yrs, with 59.41% (792) males and 40.59% (541) females. Regarding stroke lateralization, 51.02% (447) suffered a right hemisphere stroke, 48.05% (421) had a left hemisphere stroke, and 0.93% (8) presented bilateral or unspecified stroke side.

**Table 2.** Demographic and clinical characteristics of stroke survivors from the 15 included studies.

| **Study** | **Type of study** | **Fallers** | **Non-fallers** | **Age, mean (years)** | **Male** | **Female** | **Right stroke** | **Left stroke** | **Unknown/Bilateral** |
| --- | --- | --- | --- | --- | --- | --- | --- | --- | --- |
| Andersson et al. 2006, [52] | Prospective cohort | 68 | 91 | 73 | NR | NR | 70 | 85 | 4 |
| Ashburn et al. 2008, [42] | Prospective cohort | 48 | 67 | 70 | 77 | 38 | 57 | 57 | 1 |
| Beninato et al. 2009, [43] | Case series | 9 | 18 | 57 | 15 | 12 | 13 | 14 | 0 |
| Chinsongkram et al. 2014, [44] | Observational | 35 | 35 | 57 | 38 | 32 | 39 | 31 | 0 |
| Fiedorová et al. 2022, [45] | Prospective cohort | 32 | 52 | 68 | 42 | 42 | NR | NR | NR |
| Jalayondeja et al. 2014, [51] | Prospective | 25 | 72 | 62 | 59 | 38 | NR | NR | NR |
| Lee et al.<br>2016, [53] | Observational<br>cross-<br>sectional | 42 | 33 | 58 | 40 | 35 | NR | NR | NR |
| Liu et al. 2019,<br>[54] | Retrospective<br>cross-<br>sectional | 41 | 96 | 61 | 69 | 68 | 83 | 54 | 0 |
| Maeda et al.<br>2009, [46] | Retrospective<br>observational | 27 | 45 | NR | 42 | 30 | 34 | 35 | 3 |
| Persson et al.<br>2011, [55] | Prospective | 46 | 50 | 73 | 56 | 40 | 45 | 51 | 0 |
| Sahin et al.<br>2019, [47] | Cross-<br>sectional | 26 | 24 | 59 | 30 | 20 | 32 | 18 | 0 |
| Tsang et al.<br>2013, [56] | Observational | 25 | 81 | 57 | 73 | 33 | NR | NR | NR |
| Lee et al.<br>2021, [48] | Retrospective | 34 | 132 | NR | 114 | 52 | NR | NR | NR |
| Pinto et al.<br>2014, [49] | Cross-<br>sectional | 56 | 94 | 56 | 73 | 77 | 74 | 76 | 0 |
| Yang et al.<br>2016, [50] | Observational | 20 | 68 | 63 | 64 | 24 | NR | NR | NR |
| **Total** |  | **534** | **958** | **58 ± 6** | **792** | **541** | **447** | **421** | **8** |

Values represent the number of participants, unless otherwise noted; NR = Not Reported.

### Meta-analysis

A weighted random-effects model with the REML method was fitted to the 15 studies to estimate the overall effect of the Timed Up and Go test and the Berg Balance Scale on the odds of correctly identifying true non-fallers and fallers among stroke survivors. The pooled effect size indicated a negative association (OR = 0.469, 95% CI [-0.230, 1.169] (Fig. 2). However, this result was not statistically significant (*p* = 0.188). There was no evidence of heterogeneity among studies (Q = 0.797, df (20), *p* = 1.000). The estimated amount of total heterogeneity (τ^2^ = 0.000) and the I^2^ statistic (0%) indicated that none of the observed variability was due to heterogeneity between studies, suggesting consistency in the findings across the included studies.

**Figure 2.**
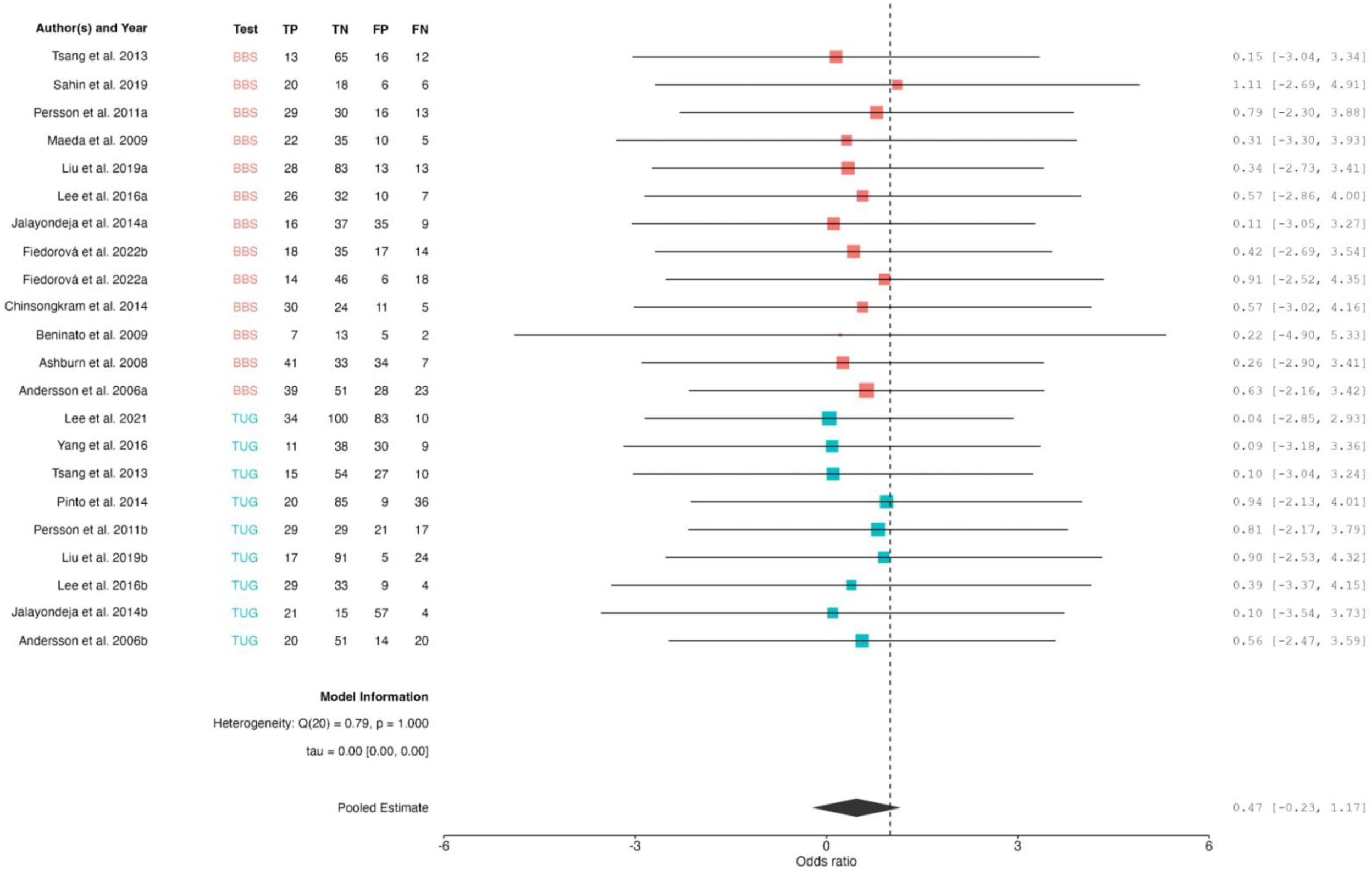
Forest plot of studies included in the meta-analysis examining the overall pooled effect of the Berg Balance Scale and Timed Up and Go test on the odds of correctly identifying true non-fallers and fallers among stroke survivors. The forest plot displays each study’s effect size (odds ratio) along with its 95% confidence interval (CI). The diamond at the bottom represents the pooled effect size association (OR = 0.469, 95% CI [-0.230, 1.169], which falls below 1, indicating a negative association between these tests and their ability to correctly identify fallers and non-fallers among stroke survivors. However, these results are not statistically significant (*p* = 0.188).

Subgroup analysis revealed that the type of tests did not significantly moderate the observed effects (Qₘ = 0.002, *p* = 0.963), indicating no significant difference in diagnostic accuracy between the Timed Up and Go test and the Berg Balance Scale. Further meta-regression analyses examining the cut-off values showed no significant moderation of diagnostic accuracy for either the Timed Up and Go test (Qₘ = 0.015, *p* = 0.902) or the Berg Balance Scale (Qₘ = 0.018, *p* = 0.894).

### Sensitivity Analysis and Risk of Bias

Publication bias assessment using Egger’s weighted test (t (20) = 0.024, *p* = 0.981), and the rank correlation test (τ = -0.030, *p* = 0.867) indicated no significant funnel plot asymmetry (Fig. 3), demonstrating the absence of publication bias in the analyzed studies. Sensitivity analysis, conducted through leave-one-out analysis, revealed that none of the studies had a significant influence on the pooled effect size.

**Figure 3.**
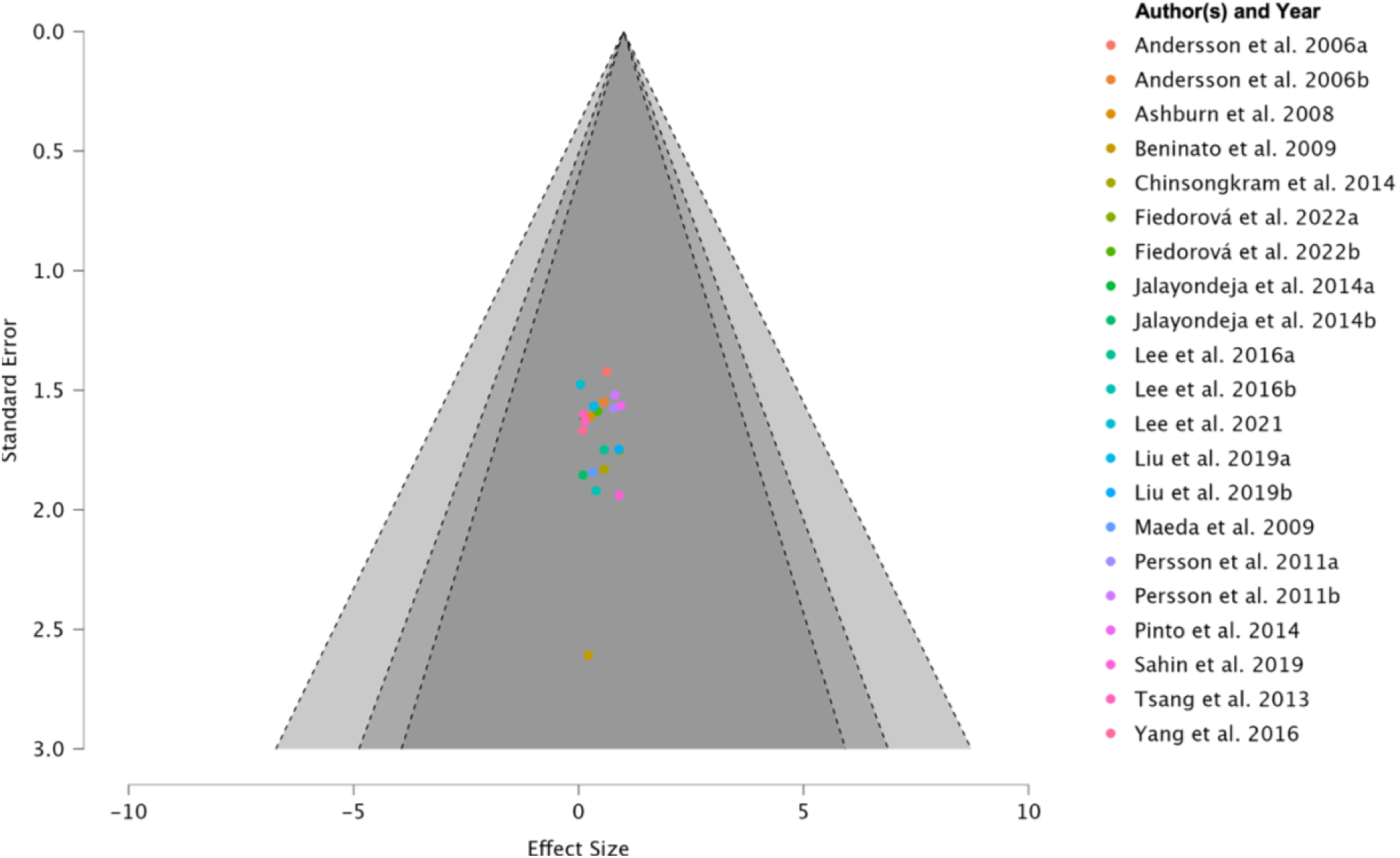
Funnel plot of the 15 studies included in the meta-analysis. Each point represents an individual study, the x-axis represents the standard error, and the y-axis is each study’s effect size (odds ratio). The symmetrical distribution of studies suggests an absence of publication bias, which is confirmed by the Egger’s weighted test (t (20) = 0.024, *p* = 0.981) and the rank correlation test (τ = -0.030, *p* = 0.867).

## DISCUSSION

This systematic review and meta-analysis evaluated the diagnostic accuracy of the most commonly used clinical assessments— the Timed Up and Go test and Berg Balance Scale—for identifying fall risk among stroke survivors. Our key findings were: 1) Over one-third (35.8%) of stroke survivors experience a fall; 2) The Timed Up and Go and the Berg Balance Scale demonstrated a negative association in accurately identifying fall risk in this population, though this was not statistically significant; 3) cut-off values varied widely across studies; however, adjusting these thresholds does not significantly improve diagnostic accuracy for fall risk classification in stroke survivors.

### Demographic and Clinical Characteristics

We analyzed data from 15 papers with a total of 1,492 stroke survivors with a mean age of 58 ± 6 years. The sex distribution (59.41% male, 40.59% female) aligns with other findings, where men generally have a higher stroke incidence at young ages (45-74 years) [57]. However, as age increases, the difference is inverted, with females experiencing a higher incidence and mortality rate above 74 years of age [57, 58]. The relatively balanced distribution of stroke lateralization (51.02% right hemisphere, 48.05% left hemisphere) reduces an overrepresentation of either hemisphere stroke. This is particularly important as hemispheric specialization influences balance and mobility. For example, a right hemisphere stroke often affects spatial awareness, left-sided neglect, and visual-spatial areas [59]. In contrast, left hemisphere strokes may have a greater impact on movement planning, sequencing, and language [60]. The fall incidence of 35.8% observed across studies is consistent with previous findings reporting fall rates between 25-73% during the first year post stroke [61–64].

### Meta-analysis

Our meta-analysis found a negative association (OR = 0.469, 95% CI [-0.230, 1.169]) between the Timed Up and Go test and the Berg Balance Scale and their diagnostic accuracy in identifying fall risk in stroke survivors. However, this finding was not statistically significant (*p* = 0.188). Nevertheless, our findings call into question their effectiveness as standard components of post-stroke clinical protocols. The clinical implications of these findings are substantial, given that clinicians currently rely on these tests to make decisions about discharge planning and stroke rehabilitation [65, 66]. If the diagnostic accuracy of these tools is limited in stroke survivors, this could directly affect patient safety and rehabilitation outcomes through inappropriate fall risk classification, potentially leading to misclassification of fall risk, resulting in a high rate of false positives and false negatives.

Several factors may explain the above negative association. First, these tests were not explicitly created for stroke survivors, but were initially developed for older adults and later applied to the stroke population [67, 68]. Even among older adults without diagnosed neurological disorders, the Timed Up and Go and the Berg Balance Scale demonstrate insufficient diagnostic accuracy to categorize fallers versus non-fallers, making them inadequate as standalone assessments for determining fall risk [23, 69]. Secondly, the complexity of balance impairment in stroke survivors extends beyond motor function. It also includes perceptual and cognitive impairments [70], which may limit the effectiveness of the Timed Up and Go and the Berg Balance Scale, as they focus only on general balance and mobility. Lastly, these tests fail to address the underlying mechanisms necessary for maintaining balance [71], specifically the contributions of the visual, vestibular, and proprioceptive systems [72]. Stroke survivors experience damage to all three sensory systems, with proprioception being affected in 11-85% of the stroke patient population [73, 74]. Therefore, without evaluating these distinct sensory systems, clinicians cannot identify the nature of the balance deficit or understand which specific postural control mechanism is compromised [75].

Our subgroup analysis found no significant difference between the Timed Up and Go test and the Berg Balance Scale in accurately categorizing fall risk (Qₘ = 0.002, *p* = 0.963). This means that clinicians face the same challenge with both tests – choosing one test over the other will not impact their diagnostic accuracy. Our meta-regression examining the cut-off values from both tests showed no significant moderation, suggesting that modifying the threshold values may not improve the ability of these tests to accurately categorize fall risk in stroke survivors. Moreover, the considerable variability in the cut-offs - ranging from 14 to 25 seconds for the Timed Up and Go test and from 19 to 50.5 for the Berg Balance Scale (see Table 1) – reflects a concerning lack of standardization across studies and represents a significant question regarding how clinicians are assessing and categorizing fall risk in stroke survivors.

It is worth noting that the absence of heterogeneity in our meta-analysis (τ^²^ = 0.000; I² = 0%) suggests consistency in these findings across the included studies. The Egger’s weighted test (t (20) = 0.024, *p* = 0.981) and the rank correlation test (τ = -0.030, *p* = 0.867) demonstrated an absence of publication bias in the included studies. Additionally, our leave-one-out sensitivity analysis, which systematically removed each study individually to evaluate its impact on the overall results, confirmed that no single study has a significant influence on the pooled effect, further strengthening the robustness of our findings.

Despite our comprehensive search using multiple databases, we identified only 15 eligible studies, which may limit the generalizability of our findings. This relatively small sample size may have affected our statistical power to detect a significant association and therefore, its interpretation requires careful consideration. Additionally, of the four most commonly used fall risk assessments, we could not assess the 6-minute walk test and 10-meter walk test because there were not enough studies that addressed their diagnostic accuracy in stroke survivors.

## CONCLUSION

In summary, this systematic review and meta-analysis found evidence of a negative association between the Timed Up and Go test and the Berg Balance Scale and their ability to accurately identify fall risk in stroke survivors. However, this finding was not statistically significant. The absence of heterogeneity across studies (I² = 0%) suggests that this finding is worth consideration despite its non-significance. Moreover, our findings raise important concerns about how these widely used tests are applied in clinical practice to categorize fall risk in stroke survivors, particularly given the considerable variability in cut-off values observed across studies, which reflects the lack of standardized assessments and cut-off values across CPGs. Future research should focus on developing or implementing a stroke-specific fall risk assessment that is objective, sensitive, and tailored to the underlying mechanisms of fall risk in stroke survivors, with appropriate cut-off values. Improving the accuracy of fall risk assessments could significantly enhance fall prevention strategies and rehabilitation care.

## Data Availability

All data produced in the present study are available upon reasonable request to the authors.

## Non-standard Abbreviations and Acronyms

CPGs: clinical practice guidelines
logOR: natural logarithm of odds ratio
OR: odds ratio
REML: restricted maximum likelihood estimator
TP: true positives
TN: true negatives
FP: false positives
FN: false negatives
TUG: Timed Up and Go
BBS: Berg Balance Scale

## ACKNOWLEDGMENTS

The authors would like to thank Mr. Santamaria Guzman, for serving as the second reviewer during the data screening process and the Auburn University Librarians for their guidance throughout with the comprehensive literature search process.

## CONFLICT OF INTEREST

All authors declare no conflict of interest.

## Notes

### Competing Interest Statement

The authors have declared no competing interest.

### Clinical Protocols

https://www.crd.york.ac.uk/PROSPERO/view/CRD420251004460

### Funding Statement

This study did not receive any funding.

### Author Declarations

This study used only openly available human data that were originally located in peer-reviewed publications indexed in databases. All data were available before the initiation of the study. These data were extracted from published articles identified through electronic searches performed in the following databases: PubMed: https://pubmed.ncbi.nlm.nih.gov/ Web of Science: https://www.webofscience.com/ SPORTDiscus (EBSCO): https://www.ebsco.com/ Ovid Medline: https://www.wolterskluwer.com/en/solutions/ovid/ovid-medline-901

